# Vaccine effectiveness of BNT162b2 against Omicron and Delta outcomes in adolescents

**DOI:** 10.1101/2022.04.07.22273319

**Authors:** Sarah A. Buchan, Lena Nguyen, Sarah E. Wilson, Sophie A. Kitchen, Jeffrey C. Kwong

## Abstract

**Introduction:** Data on vaccine effectiveness (VE) against Omicron in adolescents are limited. We estimated 2-dose and 3-dose VE against Omicron and Delta in adolescents aged 12-17 years in Ontario, Canada.

**Methods:** We conducted a test-negative design study among SARS-CoV-2-tested adolescents aged 12-17 years between November 22, 2021 (date of first Omicron detection) and March 6, 2022; we assessed Delta outcomes prior to January 2, 2022. We used multivariable logistic regression to compare the odds of vaccination in cases to symptomatic test-negative controls and calculated VE as 1-adjusted odds ratio.

**Results:** VE was lower against symptomatic Omicron infection than against Delta and decreased more rapidly over time, from 51% (95%CI, 38-61%) in the 7-59 days following a second dose to 29% (95%CI, 17-38%) after 180 days, compared to 97% (95%CI, 94-99%) and 90% (95%CI, 79-95%) for the same intervals against symptomatic Delta infection. Overall, 2-dose VE against severe outcomes caused by Omicron was 85% (95%CI, 74-91%) ≥7 days following a second dose and estimates were similar over time. VE against symptomatic Omicron infection was 62% (95%CI, 49-72%) ≥7 days following a third dose.

**Discussion:** Two-dose VE against symptomatic Omicron infection wanes over time in adolescents. While lower than observed against Delta, protection against severe outcomes appears to be maintained over time. A third dose substantially improves protection against Omicron infection, but 3-dose VE is only moderate at approximately 60% in the early period following vaccination and the duration of this protection is unknown.

## Introduction

There are limited data on vaccine effectiveness (VE) against Omicron in adolescents. In Ontario, Canada, most vaccinated adolescents completed their primary series of BNT162b2 during summer 2021; third dose eligibility (6 months following a second dose) expanded to those aged 12-17 years in February 2022. We estimated 2-dose and 3-dose VE against Omicron and Delta for this age group.

## Methods

We conducted a test-negative design among individuals aged 12-17 years and SARS-CoV-2-tested between November 22, 2021 (date of first Omicron detection) and March 6, 2022 using methods detailed elsewhere.^1,2^ We assessed Delta outcomes prior to January 2, 2022. We linked and analyzed laboratory, vaccination, reportable disease, and health administrative data for the entire eligible population at ICES. We estimated VE against symptomatic infection and severe outcomes (i.e., hospitalization or death) over time since second or third dose receipt for both Omicron and Delta, which were defined using a combination of whole genome sequencing and S-gene target failure results, and dates (Supplementary Table 1).^2^ In a sensitivity analysis, we estimated VE against symptomatic Omicron infection before and after restrictions to PCR test eligibility. We used multivariable logistic regression to compare the odds of vaccination in cases to symptomatic test-negative controls and calculated VE as 1-adjusted odds ratio.

## Ethics approval

ICES is a prescribed entity under Ontario’s Personal Health Information Protection Act (PHIPA). Section 45 of PHIPA authorizes ICES to collect personal health information, without consent, for the purpose of analysis or compiling statistical information with respect to the management of, evaluation or monitoring of, the allocation of resources to or planning for all or part of the health system. Projects that use data collected by ICES under section 45 of PHIPA, and use no other data, are exempt from REB review. The use of the data in this project is authorized under section 45 and approved by ICES’ Privacy and Legal Office.

## Results

We included 9,902 Omicron cases and 502 Delta cases (Supplementary Table 2). For the Omicron analyses, 91% of tested subjects had received two doses and 1.3% had received three doses (Supplementary Table 3). VE was lower against symptomatic Omicron infection than against Delta and decreased more rapidly over time, from 51% (95%CI, 38-61%) in the 7-59 days following a second dose to 29% (95%CI, 17-38%) after 180 days, compared to 97% (95%CI, 94-99%) and 90% (95%CI, 79-95%) for the same intervals against symptomatic Delta infection (Figure 1, Supplementary Table 4). Overall, 2-dose VE against severe outcomes due to Omicron was 85% (95%CI, 74-91%) ≥7 days following a second dose and estimates were similar over time. Although we were unable to assess 3-dose VE against severe Omicron outcomes or any Delta outcomes due to small numbers, VE against symptomatic Omicron infection was 62% (95%CI, 49-72%) ≥7 days following a third dose. Results against symptomatic Omicron infection were similar before and after changes to test eligibility criteria (Supplementary Table 5 and Supplementary Figure 1).

**Figure 1.**
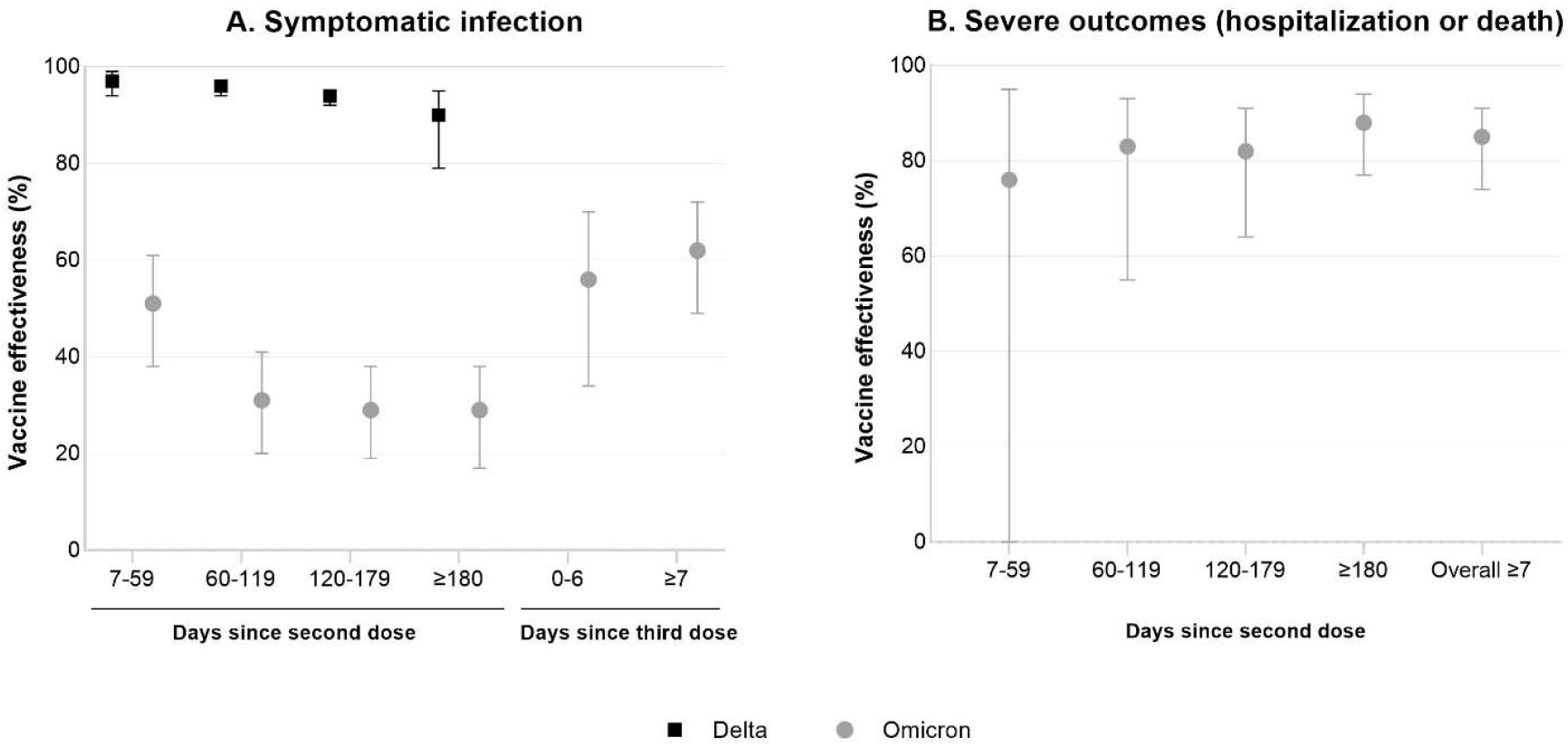
Estimates of vaccine effectiveness^a^ against symptomatic infection or severe outcomes (hospitalization or death) caused by Omicron or Delta^b^ during the period of November 22, 2021 to March 6, 2022, by time since latest dose ^a^Vaccine effectiveness estimates adjusted for: age, sex, public health unit region of residence, comorbidities, influenza vaccination status during the 2019/2020 and/or 2020/2021 influenza seasons, positive test >90 days before index date, week of testing, and neighbourhood-level information on median household income, proportion of the working population employed as non-health essential workers, mean number of persons per dwelling, and proportion of the population who self-identify as a visible minority. ^b^Delta outcomes were restricted to January 2, 2022 or prior due to small numbers and limited circulation after that date.

## Discussion

VE against symptomatic Omicron infection wanes over time in adolescents given 2-doses of BNT162b2 and was higher following a third dose. While lower than observed against Delta, protection against severe outcomes appears to be maintained over time. The declining VE against infection is similar to results from England, where VE against Omicron and Delta in adolescents aged 16-17 years decreased from 76% and 93% after 7-13 days to 23% and 84% ≥70 days post second dose, respectively.^3^ In the US, 2-dose VE against Omicron hospitalization for those aged 12-15 and 16-17 years was 92% and 94% after 14-149 days compared to 73% and 88% after ≥150 days, respectively.^4^ This difference in VE over time since second dose was not statistically significant, whereas declines in VE against emergency department (ED) visits were. VE against ED visits increased following a third dose in those aged 16-17 years.^4^ Limitations of this study included an inability to classify some specimens using SGTF or sequencing, changes to test eligibility over the study period, and potential unmeasured confounding between vaccinated and unvaccinated individuals. These results can inform third dose recommendations in adolescents, as 2-dose protection against symptomatic Omicron infection is relatively low and wanes over time, whereas protection of a second dose against severe outcomes is higher. A third dose substantially improves protection against Omicron infection in adolescents, but 3-dose VE is only moderate at approximately 60% in the early period following vaccination and the duration of this protection is unknown. Understanding the impact of a third dose on improved protection against severe outcomes will be important.

## Data Availability

The dataset from this study is held securely in coded form at ICES. While legal data sharing agreements between ICES and data providers (e.g., healthcare organizations and government) prohibit ICES from making the dataset publicly available, access may be granted to those who meet pre-specified criteria for confidential access, available at www.ices.on.ca/DAS.

## Code availability

The full dataset creation plan and underlying analytic code are available from the authors upon request, understanding that the computer programs may rely upon coding templates or macros that are unique to ICES and are therefore either inaccessible or may require modification.

## Acknowledgments

We would like to acknowledge Public Health Ontario for access to vaccination data from COVaxON, case-level data from CCM and COVID-19 laboratory data, as well as assistance with data interpretation. We also thank the staff of Ontario’s public health units who are responsible for COVID-19 case and contact management and data collection within CCM. We thank IQVIA Solutions Canada Inc. for use of their Drug Information Database. The authors are grateful to the Ontario residents without whom this research would be impossible.

## Author contributions

S.A.B. and J.C.K. designed the study. L.N. obtained the data and conducted all analyses (data set and variable creation and statistical modelling). S.A.B. and J.C.K. drafted the manuscript. All authors contributed to the analysis plan, interpreted the results, critically reviewed and edited the manuscript, approved the final version, and agreed to be accountable for all aspects of the work.

## Funding and disclaimers

This work is supported by the Applied Health Research Questions (AHRQ) Portfolio at ICES, which is funded by the Ontario Ministry of Health. For more information on AHRQ and how to submit a request, please visit www.ices.on.ca/DAS/AHRQ. This work is also supported by the Ontario Health Data Platform (OHDP), a Province of Ontario initiative to support Ontario’s ongoing response to COVID-19 and its related impacts. This work was also supported by Public Health Ontario. J.C.K. is supported by Clinician-Scientist Award from the University of Toronto Department of Family and Community Medicine.

The study sponsors did not participate in the design and conduct of the study; collection, management, analysis and interpretation of the data; preparation, review or approval of the manuscript; or the decision to submit the manuscript for publication. Parts of this material are based on data and/or information compiled and provided by the Canadian Institute for Health Information (CIHI), the Ontario Ministry of Health, and by Ontario Health (OH). Population estimates were adapted from Statistics Canada. However, the analyses, conclusions, opinions and statements expressed herein are solely those of the authors, and do not reflect those of the funding or data sources; no endorsement by ICES, MOH, MLTC, the Province of Ontario, CIHI, OH, or Statistics Canada is intended or should be inferred.

---

**Supplementary Table 1.** Number of Omicron and Delta^a^ symptomatic infections by laboratory testing categorization during the period November 22, 2021 to March 6, 2022

| **Classification criteria** | **22 NOV 2021 to**<br>**31 DEC 2021** | **1 JAN 2022 to**<br>**6 MAR 2022** |
| --- | --- | --- |
| Omicron confirmed via WGS or SGTF | 1,966 | 446 |
| Omicron classified if collection date ≥ 21 December 2021 and no or inconclusive SGTF results | 3,827 | 3,663 |
| Delta confirmed via WGS or SGTF | 447 | 0 |
| Delta classified if collection date $<$ 3 December 2021 and no or inconclusive SGTF results | 55 | 0 |

WGS: whole genome sequencing; SGTF: S-gene target failure
<sup>a</sup>Delta outcomes were restricted to January 2, 2022 or prior due to small numbers and limited circulation after that date.

**Supplementary Table 2.**
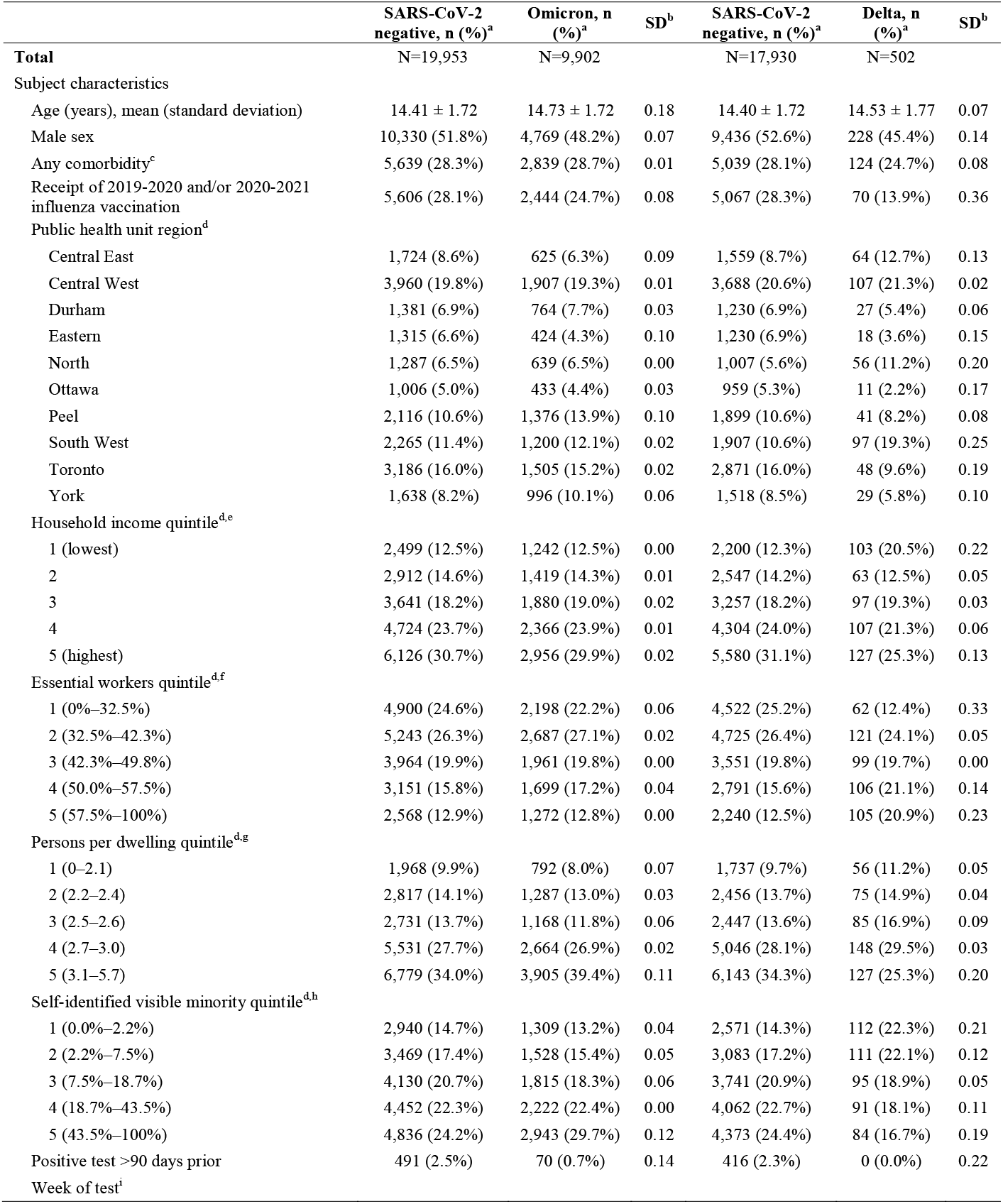
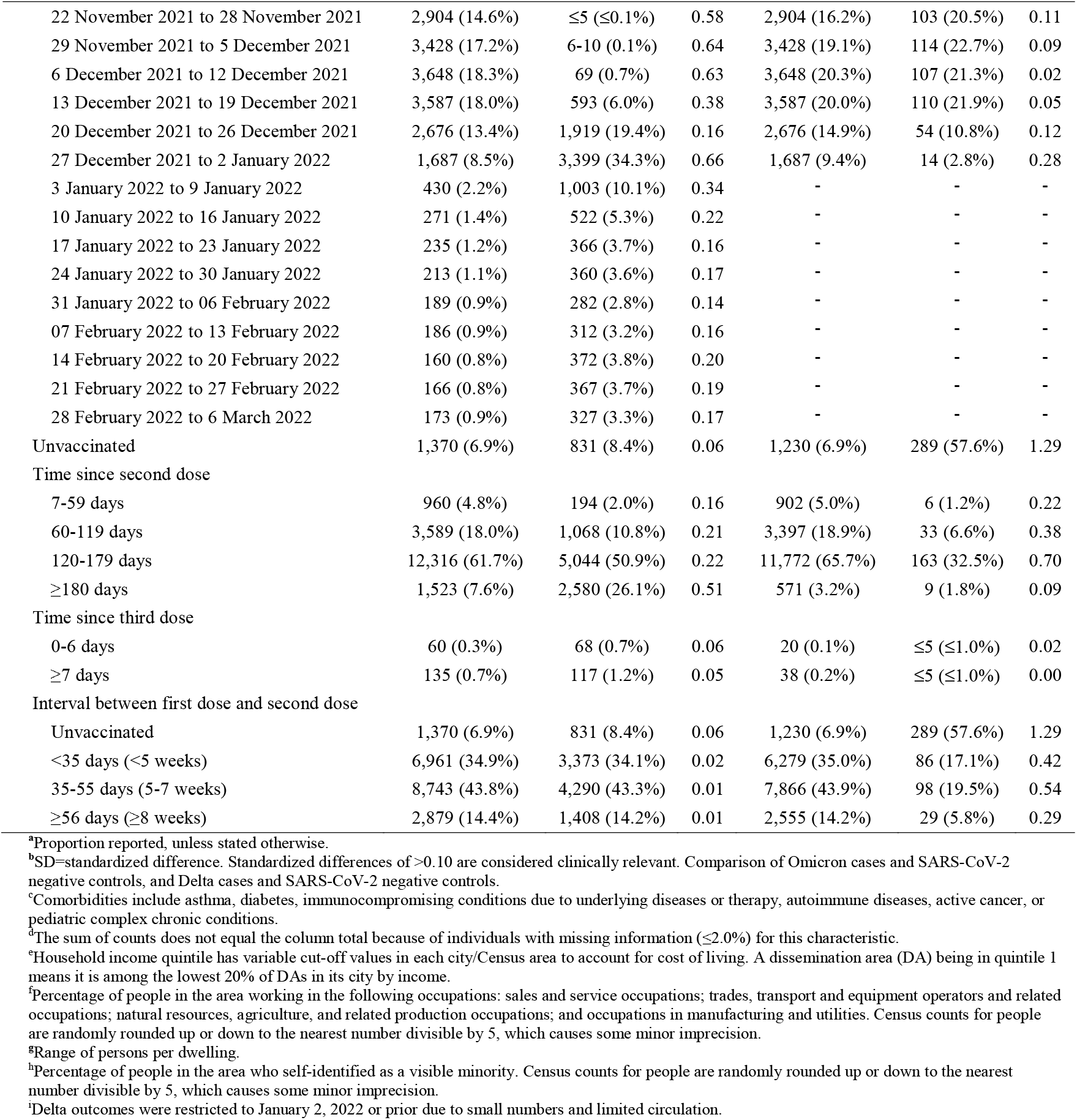
Descriptive characteristics of subjects tested for SARS-CoV-2 with COVID-19-relevant symptoms during the period November 22, 2021 to March 6, 2022 comparing Omicron cases and Delta cases with SARS-CoV-2-negative controls

**Supplementary Table 3.** Descriptive characteristics of subjects tested for SARS-CoV-2 with COVID-19-relevant symptoms during the period November 22, 2021 to March 6, 2022, comparing subjects with two doses, three doses and unvaccinated subjects, Omicron cases and SARS-CoV-2-negative controls only

|  | Unvaccinated, n (%)<sup>a</sup> | Two doses, n (%)<sup>a</sup> | SD<sup>b</sup> | Three doses, n (%)<sup>a</sup> | SD<sup>b</sup> |
| --- | --- | --- | --- | --- | --- |
| **Total** | N=2,201 | N=27,274 |  | N=380 |  |
| Subject characteristics |  |  |  |  |  |
| Age (years), mean (standard deviation) | 14.05 ± 1.80 | 14.54 ± 1.71 | 0.28 | 15.49 ± 1.58 | 0.85 |
| Male sex | 1,191 (54.1%) | 13,757 (50.4%) | 0.07 | 151 (39.7%) | 0.29 |
| Any comorbidity<sup>c</sup> | 586 (26.6%) | 7,746 (28.4%) | 0.04 | 146 (38.4%) | 0.25 |
| Receipt of 2019-2020 and/or 2020-2021 influenza vaccination | 139 (6.3%) | 7,732 (28.3%) | 0.61 | 179 (47.1%) | 1.04 |
| Public health unit region<sup>d</sup> |  |  |  |  |  |
| Central East | 201 (9.1%) | 2,115 (7.8%) | 0.05 | 33 (8.7%) | 0.02 |
| Central West | 404 (18.4%) | 5,414 (19.9%) | 0.04 | 49 (12.9%) | 0.15 |
| Durham | 132 (6.0%) | 1,982 (7.3%) | 0.05 | 31 (8.2%) | 0.08 |
| Eastern | 101 (4.6%) | 1,611 (5.9%) | 0.06 | 27 (7.1%) | 0.11 |
| North | 168 (7.6%) | 1,704 (6.2%) | 0.05 | 54 (14.2%) | 0.21 |
| Ottawa | 60 (2.7%) | 1,370 (5.0%) | 0.12 | 9 (2.4%) | 0.02 |
| Peel | 287 (13.0%) | 3,191 (11.7%) | 0.04 | 14 (3.7%) | 0.34 |
| South West | 294 (13.4%) | 3,114 (11.4%) | 0.06 | 57 (15.0%) | 0.05 |
| Toronto | 385 (17.5%) | 4,239 (15.5%) | 0.05 | 67 (17.6%) | 0.00 |
| York | 159 (7.2%) | 2,439 (8.9%) | 0.06 | 36 (9.5%) | 0.08 |
| Household income quintile<sup>d, e</sup> |  |  |  |  |  |
| 1 (lowest) | 536 (24.4%) | 3,175 (11.6%) | 0.34 | 30 (7.9%) | 0.46 |
| 2 | 396 (18.0%) | 3,900 (14.3%) | 0.10 | 35 (9.2%) | 0.26 |
| 3 | 442 (20.1%) | 5,006 (18.4%) | 0.04 | 73 (19.2%) | 0.02 |
| 4 | 401 (18.2%) | 6,589 (24.2%) | 0.15 | 100 (26.3%) | 0.20 |
| 5 (highest) | 407 (18.5%) | 8,535 (31.3%) | 0.30 | 140 (36.8%) | 0.42 |
| Essential workers quintile<sup>d, f</sup> |  |  |  |  |  |
| 1 (0%–32.5%) | 274 (12.4%) | 6,711 (24.6%) | 0.32 | 113 (29.7%) | 0.43 |
| 2 (32.5%–42.3%) | 458 (20.8%) | 7,364 (27.0%) | 0.15 | 108 (28.4%) | 0.18 |
| 3 (42.3%–49.8%) | 477 (21.7%) | 5,384 (19.7%) | 0.05 | 64 (16.8%) | 0.12 |
| 4 (50.0%–57.5%) | 479 (21.8%) | 4,315 (15.8%) | 0.15 | 56 (14.7%) | 0.18 |
| 5 (57.5%–100%) | 487 (22.1%) | 3,320 (12.2%) | 0.27 | 33 (8.7%) | 0.38 |
| Persons per dwelling quintile<sup>d, g</sup> |  |  |  |  |  |
| 1 (0–2.1) | 273 (12.4%) | 2,453 (9.0%) | 0.11 | 34 (8.9%) | 0.11 |
| 2 (2.2–2.4) | 385 (17.5%) | 3,650 (13.4%) | 0.11 | 69 (18.2%) | 0.02 |
| 3 (2.5–2.6) | 257 (11.7%) | 3,584 (13.1%) | 0.04 | 58 (15.3%) | 0.11 |
| 4 (2.7–3.0) | 573 (26.0%) | 7,517 (27.6%) | 0.03 | 105 (27.6%) | 0.04 |
| 5 (3.1–5.7) | 686 (31.2%) | 9,890 (36.3%) | 0.11 | 108 (28.4%) | 0.06 |
| Self-identified visible minority quintile<sup>d, h</sup> |  |  |  |  |  |
| 1 (0.0%–2.2%) | 356 (16.2%) | 3,820 (14.0%) | 0.06 | 73 (19.2%) | 0.08 |
| 2 (2.2%–7.5%) | 381 (17.3%) | 4,526 (16.6%) | 0.02 | 90 (23.7%) | 0.16 |
| 3 (7.5%–18.7%) | 381 (17.3%) | 5,476 (20.1%) | 0.07 | 88 (23.2%) | 0.15 |
| 4 (18.7%–43.5%) | 416 (18.9%) | 6,186 (22.7%) | 0.09 | 72 (18.9%) | 0.00 |
| 5 (43.5%–100%) | 641 (29.1%) | 7,087 (26.0%) | 0.07 | 51 (13.4%) | 0.39 |
| Positive test >90 days prior | 62 (2.8%) | 494 (1.8%) | 0.07 | ≤5 (1.3%) | 0.11 |
| Week of test |  |  |  |  |  |
| 22 November 2021 to 28 November 2021 | 248 (11.3%) | 2,657 (9.7%) | 0.05 | ≤5 (≤1.3%) | 0.49 |
| 29 November 2021 to 5 December 2021 | 273 (12.4%) | 3,162 (11.6%) | 0.02 | ≤5 (≤1.3%) | 0.50 |
| 6 December 2021 to 12 December 2021 | 292 (13.3%) | 3,421 (12.5%) | 0.02 | ≤5 (≤1.3%) | 0.49 |
| 13 December 2021 to 19 December 2021 | 251 (11.4%) | 3,917 (14.4%) | 0.09 | 12 (3.2%) | 0.32 |
| 20 December 2021 to 26 December 2021 | 229 (10.4%) | 4,337 (15.9%) | 0.16 | 29 (7.6%) | 0.10 |
| 27 December 2021 to 2 January 2022 | 416 (18.9%) | 4,631 (17.0%) | 0.05 | 39 (10.3%) | 0.25 |
| 3 January 2022 to 9 January 2022 | 135 (6.1%) | 1,282 (4.7%) | 0.06 | 16 (4.2%) | 0.09 |
| 10 January 2022 to 16 January 2022 | 89 (4.0%) | 681 (2.5%) | 0.09 | 23 (6.1%) | 0.09 |
| 17 January 2022 to 23 January 2022 | 56 (2.5%) | 535 (2.0%) | 0.04 | 10 (2.6%) | 0.01 |
| 24 January 2022 to 30 January 2022 | 48 (2.2%) | 512 (1.9%) | 0.02 | 13 (3.4%) | 0.08 |
| 31 January 2022 to 06 February 2022 | 58 (2.6%) | 399 (1.5%) | 0.08 | 14 (3.7%) | 0.06 |
| 07 February 2022 to 13 February 2022 | 40 (1.8%) | 435 (1.6%) | 0.02 | 23 (6.1%) | 0.22 |
| 14 February 2022 to 20 February 2022 | 25 (1.1%) | 483 (1.8%) | 0.05 | 24 (6.3%) | 0.28 |
| 21 February 2022 to 27 February 2022 | 21 (1.0%) | 446 (1.6%) | 0.06 | 66 (17.4%) | 0.59 |
| 28 February 2022 to 6 March 2022 | 20 (0.9%) | 376 (1.4%) | 0.04 | 104 (27.4%) | 0.82 |
| Unvaccinated | 2,201 (100%) | 0 (0.0%) | - | 0 (0.0%) | - |
| Time since second dose |  |  |  |  |  |
| 7-59 days | 0 (0.0%) | 1,154 (4.2%) | 0.30 | 0 (0.0%) | - |
| 60-119 days | 0 (0.0%) | 4,657 (17.1%) | 0.64 | 0 (0.0%) | - |
| 120-179 days | 0 (0.0%) | 17,360 (63.7%) | 1.87 | 0 (0.0%) | - |
| ≥180 days | 0 (0.0%) | 4,103 (15.0%) | 0.60 | 0 (0.0%) | - |
| Time since third dose |  |  |  |  |  |
| 0-6 days | 0 (0.0%) | 0 (0.0%) | - | 128 (33.7%) | 1.01 |
| ≥7 days | 0 (0.0%) | 0 (0.0%) | - | 252 (66.3%) | 1.98 |
| Interval between dose 1 and dose 2 |  |  |  |  |  |
| Unvaccinated | 2,201 (100%) | 0 (0.0%) | - | 0 (0.0%) | - |
| <35 days (<5 weeks) | 0 (0.0%) | 10,187 (37.4%) | 1.09 | 147 (38.7%) | 1.12 |
| 35-55 days (5-7 weeks) | 0 (0.0%) | 12,859 (47.1%) | 1.34 | 174 (45.8%) | 1.30 |
| ≥56 days (≥8 weeks) | 0 (0.0%) | 4,228 (15.5%) | 0.61 | 59 (15.5%) | 0.61 |

<sup>a</sup>Proportion reported, unless stated otherwise.
<sup>b</sup>SD=standardized difference. Standardized differences of >0.10 are considered clinically relevant. Comparison of subjects who have received 2 or 3 doses with unvaccinated subjects.
<sup>c</sup>Comorbidities include asthma, diabetes, immunocompromising conditions due to underlying diseases or therapy, autoimmune diseases, active cancer, or pediatric complex chronic conditions.
<sup>d</sup>The sum of counts does not equal the column total because of individuals with missing information (≤1.0%) for this characteristic.
<sup>e</sup>Household income quintile has variable cut-off values in each city/Census area to account for cost of living. A dissemination area (DA) being in quintile 1 means it is among the lowest 20% of DAs in its city by income.
<sup>f</sup>Percentage of people in the area working in the following occupations: sales and service occupations; trades, transport and equipment operators and related occupations; natural resources, agriculture, and related production occupations; and occupations in manufacturing and utilities. Census counts for people are randomly rounded up or down to the nearest number divisible by 5, which causes some minor imprecision.
<sup>g</sup>Range of persons per dwelling.
<sup>h</sup>Percentage of people in the area who self-identified as a visible minority. Census counts for people are randomly rounded up or down to the nearest number divisible by 5, which causes some minor imprecision.

**Supplementary Table 4.** Estimates of vaccine effectiveness^a^ against symptomatic infection or severe outcomes (hospitalization or death) caused by Omicron or Delta^b^ during the period November 22, 2021 to March 06, 2022, by time since latest dose

| Outcome | Dose number | Days since latest dose | SARS-CoV-2 negative controls, n | Omicron-positive cases, n | Vaccine effectiveness against Omicron (95% CI) | SARS-CoV-2 negative controls, n<sup>b</sup> | Delta-positive cases, n<sup>b</sup> | Vaccine effectiveness against Delta (95% CI)<sup>b</sup> |
| --- | --- | --- | --- | --- | --- | --- | --- | --- |
| Symptomatic infection | Second dose | 7-59 days | 960 | 194 | 51 (38, 61) | 902 | 6 | 97 (94, 99) |
|  |  | 60-119 days | 3,589 | 1,068 | 31 (20, 41) | 3,397 | 33 | 96 (94, 97) |
|  |  | 120-179 days | 12,316 | 5,044 | 29 (19, 38) | 11,772 | 163 | 94 (92, 95) |
|  |  | ≥180 days | 1,523 | 2,580 | 29 (17, 38) | 571 | 9 | 90 (79, 95) |
|  | Third dose | 0-6 days | 60 | 68 | 56 (34, 70) | N/A | N/A | N/A<sup>c</sup> |
|  |  | ≥7 days | 135 | 117 | 62 (49, 72) | N/A | N/A | N/A<sup>c</sup> |
| Severe Outcomes | Second dose | 7-59 days | 960 | ≤5 | 76 (-10, 95) | 902 | 0 | <sup>d</sup> |
|  |  | 60-119 days | 3,589 | 4-8 | 83 (55, 93) | 3,397 | 0 | <sup>d</sup> |
|  |  | 120-179 days | 12,316 | 18 | 82 (64, 91) | 11,772 | 0 | <sup>d</sup> |
|  |  | ≥180 days | 1,523 | 22 | 88 (77, 94) | 571 | 0 | <sup>d</sup> |
|  |  | Overall ≥7 days | 18,388 | 49 | 85 (74, 91) | 16,642 | 0 | <sup>d</sup> |

<sup>a</sup>Vaccine effectiveness estimates adjusted for: age, sex, public health unit region of residence, comorbidities, influenza vaccination status during the 2019/2020 and/or 2020/2021 influenza seasons, positive test >90 days before index date, week of testing, and neighbourhood-level information on median household income, proportion of the working population employed as non-health essential workers, mean number of persons per dwelling, and proportion of the population who self-identify as a visible minority.
<sup>b</sup>Estimated to January 2, 2022 only due to small numbers and limited circulation after that date.
<sup>c</sup>Not estimated
<sup>d</sup>Vaccine effectiveness estimated as 100% based on zero vaccinated test-positive cases.

**Supplementary Table 5.** Characteristics of subjects tested for SARS-CoV-2 with COVID-19-relevant symptoms during the period November 22, 2021 to March 6, 2022, before and after PCR testing restrictions, Omicron cases and SARS-CoV-2-negative controls only

|  | Tested on or before<br>December 31, 2021<sup>a</sup> | Tested after<br>December 31, 2021<sup>a</sup> | SD<sup>b</sup> |
| --- | --- | --- | --- |
| **Total** | N=23,615 | N=6,240 |  |
| Subject characteristics |  |  |  |
| Age (years), mean (standard deviation) | 14.49 ± 1.73 | 14.60 ± 1.73 | 0.06 |
| Male sex | 12,247 (51.9%) | 2,852 (45.7%) | 0.12 |
| Any comorbidity | 6,671 (28.2%) | 1,807 (29.0%) | 0.02 |
| Receipt of 2019-2020 and/or 2020-2021<br>influenza vaccination | 6,489 (27.5%) | 1,561 (25.0%) | 0.06 |
| Public health unit region<sup>d</sup> |  |  |  |
| Central East | 1,955 (8.3%) | 394 (6.3%) | 0.08 |
| Central West | 4,965 (21.0%) | 902 (14.5%) | 0.17 |
| Durham | 1,630 (6.9%) | 515 (8.3%) | 0.05 |
| Eastern | 1,448 (6.1%) | 291 (4.7%) | 0.07 |
| North | 1,158 (4.9%) | 768 (12.3%) | 0.27 |
| Ottawa | 1,262 (5.3%) | 177 (2.8%) | 0.13 |
| Peel | 2,741 (11.6%) | 751 (12.0%) | 0.01 |
| South West | 2,403 (10.2%) | 1,062 (17.0%) | 0.20 |
| Toronto | 3,800 (16.1%) | 891 (14.3%) | 0.05 |
| York | 2,170 (9.2%) | 464 (7.4%) | 0.06 |
| Household income quintile<sup>d,e</sup> |  |  |  |
| 1 (lowest) | 2,787 (11.8%) | 954 (15.3%) | 0.10 |
| 2 | 3,320 (14.1%) | 1,011 (16.2%) | 0.06 |
| 3 | 4,311 (18.3%) | 1,210 (19.4%) | 0.03 |
| 4 | 5,742 (24.3%) | 1,348 (21.6%) | 0.06 |
| 5 (highest) | 7,390 (31.3%) | 1,692 (27.1%) | 0.09 |
| Essential workers quintile<sup>d,f</sup> |  |  |  |
| 1 (0%–32.5%) | 5,912 (25.0%) | 1,186 (19.0%) | 0.15 |
| 2 (32.5%–42.3%) | 6,400 (27.1%) | 1,530 (24.5%) | 0.06 |
| 3 (42.3%–49.8%) | 4,610 (19.5%) | 1,315 (21.1%) | 0.04 |
| 4 (50.0%–57.5%) | 3,673 (15.6%) | 1,177 (18.9%) | 0.09 |
| 5 (57.5%–100%) | 2,874 (12.2%) | 966 (15.5%) | 0.10 |
| Persons per dwelling quintile<sup>d,g</sup> |  |  |  |
| 1 (0–2.1) | 2,136 (9.0%) | 624 (10.0%) | 0.03 |
| 2 (2.2–2.4) | 3,104 (13.1%) | 1,000 (16.0%) | 0.08 |
| 3 (2.5–2.6) | 3,084 (13.1%) | 815 (13.1%) | 0.00 |
| 4 (2.7–3.0) | 6,585 (27.9%) | 1,610 (25.8%) | 0.05 |
| 5 (3.1–5.7) | 8,559 (36.2%) | 2,125 (34.1%) | 0.05 |
| Self-identified visible minority quintile<sup>d,h</sup> |  |  |  |
| 1 (0.0%–2.2%) | 3,165 (13.4%) | 1,084 (17.4%) | 0.11 |
| 2 (2.2%–7.5%) | 3,894 (16.5%) | 1,103 (17.7%) | 0.03 |
| 3 (7.5%–18.7%) | 4,821 (20.4%) | 1,124 (18.0%) | 0.06 |
| 4 (18.7%–43.5%) | 5,425 (23.0%) | 1,249 (20.0%) | 0.07 |
| 5 (43.5%–100%) | 6,165 (26.1%) | 1,614 (25.9%) | 0.01 |
| Positive test >90 days prior | 450 (1.9%) | 111 (1.8%) | 0.01 |
| Week of test |  |  |  |
| 22 November 2021 to 28 November 2021 | 2,906 (12.3%) | 0 (0.0%) | 0.53 |
| 29 November 2021 to 5 December 2021 | 3,437 (14.6%) | 0 (0.0%) | 0.58 |
| 6 December 2021 to 12 December 2021 | 3,717 (15.7%) | 0 (0.0%) | 0.61 |
| 13 December 2021 to 19 December 2021 | 4,180 (17.7%) | 0 (0.0%) | 0.66 |
| 20 December 2021 to 26 December 2021 | 4,595 (19.5%) | 0 (0.0%) | 0.70 |
| 27 December 2021 to 2 January 2022 | 4,780 (20.2%) | 306 (4.9%) | 0.48 |
| 3 January 2022 to 9 January 2022 | 0 (0.0%) | 1,433 (23.0%) | 0.77 |
| 10 January 2022 to 16 January 2022 | 0 (0.0%) | 793 (12.7%) | 0.54 |
| 17 January 2022 to 23 January 2022 | 0 (0.0%) | 601 (9.6%) | 0.46 |
| 24 January 2022 to 30 January 2022 | 0 (0.0%) | 573 (9.2%) | 0.45 |
| 31 January 2022 to 6 February 2022 | 0 (0.0%) | 471 (7.5%) | 0.40 |
| 07 February 2022 to 13 February 2022 | 0 (0.0%) | 498 (8.0%) | 0.42 |
| 14 February 2022 to 20 February 2022 | 0 (0.0%) | 532 (8.5%) | 0.43 |
| 21 February 2022 to 27 February 2022 | 0 (0.0%) | 533 (8.5%) | 0.43 |
| 28 February 2022 to 6 March 2022 | 0 (0.0%) | 500 (8.0%) | 0.42 |
| Unvaccinated | 1,675 (7.1%) | 526 (8.4%) | 0.05 |
| Time since second dose |  |  |  |
| 7-59 days | 1,004 (4.3%) | 150 (2.4%) | 0.10 |
| 60-119 days | 4,126 (17.5%) | 531 (8.5%) | 0.27 |
| 120-179 days | 15,499 (65.6%) | 1,861 (29.8%) | 0.77 |
| ≥180 days | 1,227 (5.2%) | 2,876 (46.1%) | 1.06 |
| Time since third dose |  |  |  |
| 0-6 days | 32 (0.1%) | 96 (1.5%) | 0.15 |
| ≥7 days | 52 (0.2%) | 200 (3.2%) | 0.23 |
| Interval between first dose and second dose |  |  |  |
| Unvaccinated | 1,675 (7.1%) | 526 (8.4%) | 0.05 |
| <35 days (<5 weeks) | 8,249 (34.9%) | 2,085 (33.4%) | 0.03 |
| 35-55 days (5-7 weeks) | 10,371 (43.9%) | 2,662 (42.7%) | 0.03 |
| ≥56 days (≥8 weeks) | 3,320 (14.1%) | 967 (15.5%) | 0.04 |

<sup>a</sup>Proportion reported, unless stated otherwise.
<sup>b</sup>SD=standardized difference. Standardized differences of >0.10 are considered clinically relevant. Comparison of subjects tested after December 31, 2021 and before December 31, 2021.
<sup>c</sup>Comorbidities include asthma, diabetes, immunocompromising conditions due to underlying diseases or therapy, autoimmune diseases, active cancer, or pediatric complex chronic conditions
<sup>d</sup>The sum of counts does not equal the column total because of individuals with missing information (≤1.1%) for this characteristic.
<sup>e</sup>Household income quintile has variable cut-off values in each city/Census area to account for cost of living. A dissemination area (DA) being in quintile 1 means it is among the lowest 20% of DAs in its city by income.
<sup>f</sup>Percentage of people in the area working in the following occupations: sales and service occupations; trades, transport and equipment operators and related occupations; natural resources, agriculture, and related production occupations; and occupations in manufacturing and utilities.
Census counts for people are randomly rounded up or down to the nearest number divisible by 5, which causes some minor imprecision.
<sup>g</sup>Range of persons per dwelling.
<sup>h</sup>Percentage of people in the area who self-identified as a visible minority. Census counts for people are randomly rounded up or down to the nearest number divisible by 5, which causes some minor imprecision.

**Supplementary Figure 1.**
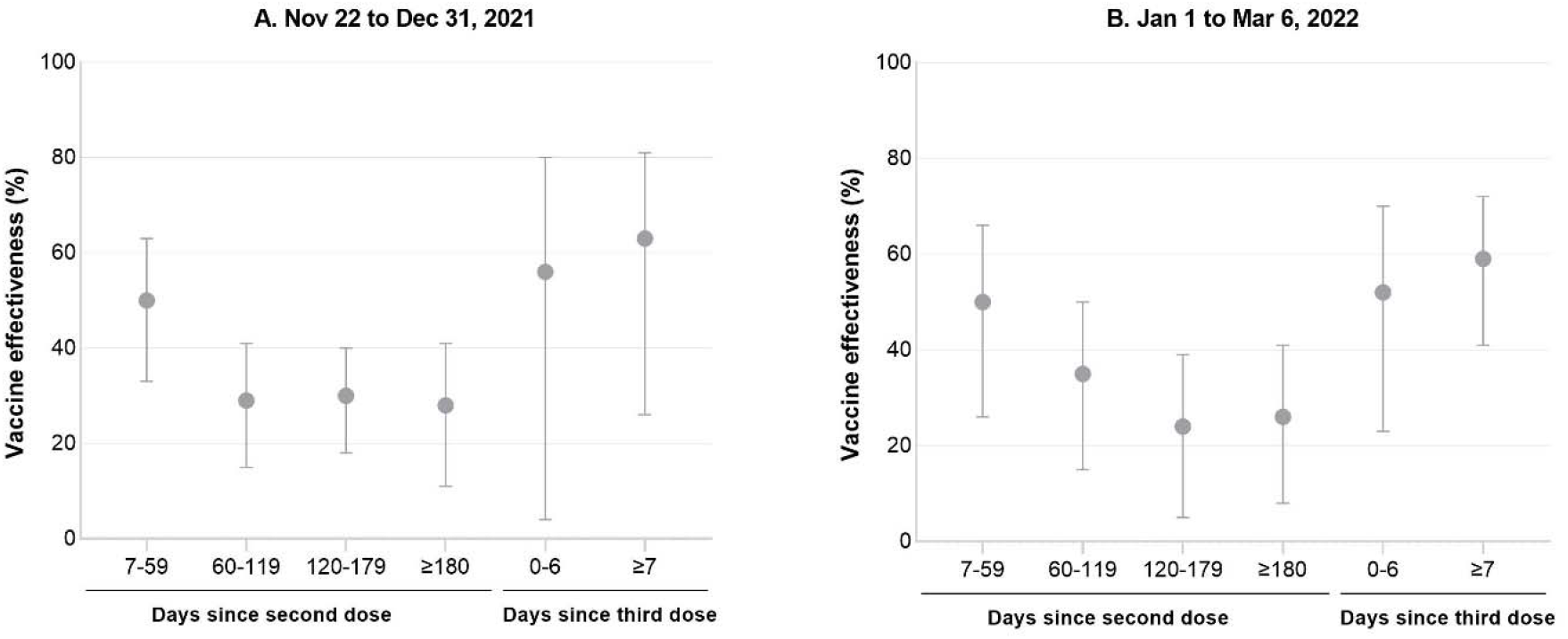
Estimates of vaccine effectiveness^a^ against symptomatic infection caused by Omicron during the period November 22, 2021 to March 6, 2022, by time since latest dose, before and after PCR testing restrictions ^a^Vaccine effectiveness estimates adjusted for: age, sex, public health unit region of residence, comorbidities, influenza vaccination status during the 2019/2020 and/or 2020/2021 influenza seasons, positive test >90 days before index date, week of testing, and neighbourhood-level information on median household income, proportion of the working population employed as non-health essential workers, mean number of persons per dwelling, and proportion of the population who self-identify as a visible minority.

